# Brainstem-cortical dynamics of Parkinson’s disease autonomic failure

**DOI:** 10.64898/2026.05.07.26352636

**Authors:** Herberto Dhanis, Charlotte Dore, Nicola Smith, Gabriel Sheehan, Pranay Yadav, Christian Lambert

## Abstract

In Parkinson’s disease (PD), failure to regulate blood pressure is a common comorbidity that is hard to treat, worsens quality-of-life and often precedes diagnosis. It is caused by α-synuclein-mediated damage to the autonomic nervous system (ANS) that precedes overt symptoms, and while central dysfunction is a key component it remains uncharacterised *in-vivo* due to various imaging challenges. Here, we characterised the function of a brainstem-cortical ANS network in relation to cardiovascular autonomic failure.

81 individuals with early-PD and 65 aged-matched healthy controls (HC) performed resting-state 3T fMRI, neurological assessments and a 3-minute standing test: lied down for 5-minutes, had blood-pressure measured, then stood-up and had blood-pressure measured every minute for 3-minutes. We used Dynamic Causal Modelling to investigate how drops in blood pressure linked to effective connectivity in medullary and relay nuclei, hypothalamus, and cortical autonomic regions.

Steep drops in blood-pressure in PD were associated with an overall inhibitory pattern departing from the rostro-ventrolateral medulla, a region responsible for sympathetic-mediated increase in blood pressure, as well as increased self-inhibition of this region. Additionally, the patient model was dominated by bottom-up connectivity from medullary regions to relay and cortical ones, potentially pertaining to state-signalling to higher-order regions, given the decreased ability of medullary regions to regulate blood pressure. Conversely, in HC, we observed widespread and unspecific weakening of connectivity.

Our findings elucidate for the first time how a baseline dysfunction originating from the medulla, may cause poor central ANS reactivity to orthostatic stimuli and predispose those with PD to drops in blood pressure upon standing. These findings are compatible with known pathophysiology literature, pave the way to more in-depth characterisations of central autonomic dysfunction *in-vivo*, and may result in functional signatures useful to track disease progression or outcomes in clinical trials.

## Introduction

Autonomic failure is one of the most common non-motor features of Parkinson’s disease (PD) (Mendoza-Velásquez et al., 2019; Schapira et al., 2017). It may precede the diagnosis of PD in up to a quarter of cases by years or sometimes decades (J.-A. Palma & Kaufmann, 2014; Postuma et al., 2013), it worsens as disease progresses (Stanković et al., 2019; Stewart et al., 2023) and is associated with poor outcomes including more rapid progression to dementia (Martinez-Nunez et al., 2026) and lower survival rates (De Pablo-Fernandez et al., 2017). Common clinical presentations of autonomic failure include gastrointestinal issues such as constipation (Cersosimo & Benarroch, 2008), swallowing difficulties (Pfeiffer, 2018), urogenital dysfunction (Allan, 2019; Winge & Fowler, 2006) and neurogenic orthostatic hypotension (Mendoza-Velásquez et al., 2019).

Neurogenic orthostatic hypotension (nOH) is defined as a sustained decrease in blood pressure upon standing, typically at least 20 mmHg systolic or 10 mmHg diastolic within 3 minutes of standing (although these cut-off values vary slightly depending on the context), that is caused by dysfunction at the level of the autonomic nervous system (Jain & Goldstein, 2012; Mendoza-Velásquez et al., 2019). In Parkinson’s, manifestations of nOH can range from asymptomatic or very mild postural symptoms such as light-headiness or dizziness, through to extremely disabling syncopal episodes. These worsen quality-of-life and significantly increase the risk of falls and can result severe, life changing injuries (Idiaquez & Roman, 2011). Moreover, nOH is likely to contribute to other PD-related non-motor symptoms, such as fatigue, poor concentration and cognitive decline (Loureiro et al., 2023; Martinez-Nunez et al., 2026), that may in part be due to associated cerebral hypoperfusion (Kenny et al., 2002).

In PD, autonomic failure is caused by abnormal accumulation of misfolded α-synuclein proteins in neurons of the autonomic nervous system (Wakabayashi, 2020; Wakabayashi & Takahashi, 1997). A key component to nOH is peripheral noradrenergic denervation of the heart and central baroreflex failure (Goldstein & Sharabi, 2019; Jain & Goldstein, 2012; Lamotte et al., 2020). The specific role of central structures remains less understood, but it is highly likely that these also contribute to the pathophysiology. Damage to medullary sympathoexcitatory groups responsible for central regulation of blood pressure, such as the rostral ventrolateral medulla (RVLM) (Benarroch, 2004; ‘Central Control of Autonomic Function and Involvement in Neurodegenerative Disorders’, 2013; Coon et al., 2018; Guyenet, 2006), and cardiovascular control, such as the nucleus of the solitary tract and locus coeruleus (Braak et al., 2003), is seen early in the disease course and even in the absence of damage to peripheral sympathetic nerves in the skin and muscles (Krämer et al., 2019). Abnormal α-synuclein aggregates are found in the medulla at all stages of PD (Andersen et al., 2025; Braak et al., 2003), with post-mortem studies showing involvement during of structures such as the medullary sympathetic (Benarroch, 2004), vagal nuclei (Benarroch et al., 2006) and other important relay and integrative nuclei of the autonomic nervous system, such as the medial parabrachial nuclei (MPB) and the periacqueductal grey (PAG) (Seidel et al., 2015).

Investigating the central autonomic nervous system has mostly relied on animal (Guyenet, 2006; Joers et al., 2014; Van Den Berge et al., 2021) and post-mortem human studies (Dickson et al., 2008; Goldstein, Sullivan, et al., 2011; Seidel et al., 2015) due to the considerable technological obstacles in imaging the small nuclei of the human brainstem (Beissner, 2015; Sclocco et al., 2018). Consequently, the central mechanisms of autonomic failure remain poorly understood, which represents a critical gap. Better defining the central mechanisms of nOH *in-vivo* would not only help understand how autonomic nervous system function changes and adapts in neurodegenerative disorders, but help clarify how these changes contribute to other features of Parkinson’s such as fatigue, mood disorders and cognitive dysfunction. Furthermore it may also help identify early biomarkers of disease, given that neuronal dysfunction due to α-synuclein deposition occurs long before neuronal loss (Jellinger, 2012; Yaribash et al., 2025), and atrophy only becomes relevant once significant and irreversible neuronal loss has occurred (Burke & O’Malley, 2013; Ma et al., 1997).

Recent neuroimaging developments in functional MRI (fMRI) have shown that it is possible to reliably and precisely define *functional connectivity*, that is the statistical dependencies, between the brainstem and cortex (Cauzzo et al., 2022; Hansen et al., 2024; Singh et al., 2022). These studies marked a crucial advance in identifying relationships between different brainstem and cortical functional networks, such as preferential connectivity between groups of brainstem nuclei (i.e. PAG, red nucleus, median raphe) with inferior frontal cortical regions, or of other groups (i.e. locus coeruleus, MPB) with anterior limbic regions (Hansen et al., 2024). The limitation with this approach is it is unable to disambiguate the directionality of these relationships between different cortico-brainstem circuits. For the study of central autonomic function this is particularly important, as brainstem nuclei are specific in their function and projections (e.g. the nucleus of the solitary tract is an afferent receiver and integrator which relays signal to other nuclei, RVLM is specifically a sympathetic efferent), and changes in the directionality with cortical networks have inherently different meaning, with brainstem-to-cortex connectivity conveying signals that provide an update on the current physiological state of the body (“state-setting”, e.g.(Berntson & Khalsa, 2021; Gut & Winn, 2016)) whereas cortex-to-brainstem connectivity may provide feedback that can adapt and modulate the incoming physiological signals (“gain-setting”, e.g.(Colivicchi et al., 2004; Elston & Bilkey, 2017)).

To address this, here we applied an advanced Bayesian framework (K. J. Friston et al., 2014) to estimate the *effective* connectivity, the causal directed influence of different neural populations, along a brainstem-cortical autonomic network. An important difference from other approaches is that this is a generative framework. Data is modelled under several assumptions, such as: predefined neurovascular and haemodynamic parameters so that the predicted data follows observable brain properties (K. J. Friston et al., 2000); parameters that specify strength and directionality of neuronal connectivity; and finally parameters that define the expected measurement noise for fMRI settings (K. J. Friston et al., 2003). A key advantage of this framework, beyond the ability to estimate causal directional influence between brain regions, is that once many models of neuronal connectivity have been estimated, their model evidence can be compared to infer which model is most likely to account for the observed data. Ultimately, it means we can retrieve both the common excitatory or inhibitory nature of the connections across all individuals, but also how neuronal connections become weaker or stronger with population-level covariates (Zeidman, Jafarian, Seghier, et al., 2019).

This work focuses on a cohort with early-stage PD, defined as less than two-years following motor diagnosis. Given α-synuclein pathology is present within autonomic nuclei of the brainstem from the earliest stages of PD, even in those without any overt signs of autonomic dysfunction (Andersen et al., 2025; Braak et al., 2003), our hypothesis was that pathological changes in connectivity of the central regulatory autonomic nervous system would already be present. Specifically, we hypothesised that variation in the orthostatic response of blood-pressure upon standing in Parkinson’s would be linked to changes in effective connectivity in the cardiovascular medullary regions. We used baseline data from healthy controls (HC) and recently diagnosed PD from an ongoing longitudinal study in Parkinson’s. Using resting state fMRI data combined with an *a priori* autonomic network composed of brainstem, hypothalamic nuclei and cortical regions relevant for blood-pressure autoregulation, our aims were to:

1. Establish the baseline autonomic network architecture in PD.
2. Characterise how functional alterations in these regions (i.e. altered connectivity) relates to drops in blood-pressure upon standing in PD.
3. Define how both baseline and changes compare to aged-matched healthy individuals.

## Materials and methods

### Participants

We used baseline data from 81 individuals with early PD and 65 aged-matched healthy controls (HC) that were taking part in the ongoing “*Quantitative MRI for anatomical phenotyping in Parkinson’s disease*” project (QMAP-PD) and had resting-state fMRI available. All enrolled between October 2018 and August 2022, were aged between 30 and 75 years, with no contraindications to MRI scanning and no other major neurological diagnoses. Written informed consent was obtained for each participant upon arrival. The study was approved by London - Fulham Research Ethics Committee (R&D / Sponsor Reference Number(s): 18/0232, Study Registration Number: IRAS number 247033).

### Clinical and Autonomic Assessments

All study participants went through detailed medical and neurological examinations with the same consultant neurologist specialised in movement disorders (CL). This included all four parts of the Movement Disorder Society Unified Parkinson’s disease Rating Scale (MDS-UPDRS)(Goetz et al., 2008), which assessed non-motor experiences, motor-experiences, motor evaluation (i.e. severity of motor symptoms), and motor complication with treatment, respectively, as well as the Montreal Cognitive Assessment (MoCA)(Nasreddine et al., 2005) to assess cognitive function. Additionally, all participants completed specific autonomic questionnaires, including the Scale for Outcomes in Parkinson’s disease – Autonomic Dysfunction (SCOPA-AUT) (Visser et al., 2004), the Sialorrhea Clinical Scale for Parkinson’ disease (SCS-PD) (Perez Lloret et al., 2007), and the self-completed Non-Motor Symptom Questionnaire (NMSQ) (Chaudhuri et al., 2006) that includes many questions on autonomic dysfunction. Participants also performed a 3-minute orthostatic challenge: individuals laid down for 5-minutes with their blood pressure being checked at the end, then stood for 3-minutes and blood pressure was checked every minute.

### Imaging

Participants completed a 5-minute resting-state fMRI acquisition at the Functional Imaging Laboratory in Queen Square (London, UK), on a Siemens MAGNETOM Prisma 3T scanner that used a 64-channel head-and-neck coil. Echo-planar sequences were used (EPI, TR = 1.18s, TE = 32.0ms, flip-angle = 64°, multi-band acceleration factor = 6), with an in-plane resolution of 2.0 x 2.0 mm (104 x 90), a slice thickness of 2.0mm (no gaps, 78 slices, FOV = 208mm) and slices were aligned to the bi-commisural plan with the acquisition volume aligned to cover the entire brain hemispheres and brainstem. Participants were instructed to keep their eyes closed and not fall asleep, and whilst they did this all the lights and projector in the scanner were turned off. With regards to dopaminergic medication, all participants attended their baseline assessment in the clinically effective off state having withheld this overnight. They were allowed to take these immediately before entering the scanner environment, and the rsFMRI was the first acquisition performed (time from medication to scan <10 minutes).

Structural acquisitions were acquired via the multiple parametric mapping (MPM) protocol (for the full list of parameters consult Callaghan and colleagues 2019(Callaghan et al., 2019)). This includes B1 field (B1-map) using the 3D EPI SE/STE method(Lutti et al., 2010, 2012) and B0-field map to correct for inhomogeneities, followed by 3D multi-echo, fast low angle shot (FLASH) scans with T1, proton density or MT weighting. The total scanning time for MPM acquisition was 27 mins and 14 seconds.

### Preprocessing

#### Structural MRI

The MPM data was pre-processed using the hMRI toolbox (Tabelow et al., 2019). The default tissue probability maps for the hMRI toolbox were changed to use the brain-spine priors that allowed us to include more of the cervical spinal cord present in the original images. This step required the proton density bias correction to be only based on grey and white matter segmentations, instead of brain plus soft tissue, because of the change in the distribution of non-brain tissue due to wrap. All weighted images were segmented using SPM12 and bias corrected using 20mm FWHM. Only MTw maps were used for this work.

#### Functional MRI

fMRI resting-state data was pre-processed using SPM12 (Functional Imaging Laboratory, UCL). The first six volumes were removed so steady state could be achieved. We then performed slice-time acquisition correction and realignment with simultaneous correction of field inhomogeneities. An intermediate step used the ART toolbox (Gabrielli lab, MIT) to identify outliers with a framewise displacement above 0.9mm and recompute the mean without those volumes. Following this, structural images were co-registered to the mean functional image and segmented to obtain deformation fields to MNI space. Indirect normalization to MNI space was performed, and liberal spatial smoothing was used (FWHM = 4mm), considering our interest in the brainstem. Data was then denoised using an independent component analysis approach for automatic removal of artifacts, optimized for the removal of movement noise (Pruim et al., 2015). Finally, we regressed out five principal components extracted from a brainstem CSF mask in order to reduce the influence of physiological noise (akin to anatomical component correction (Behzadi et al., 2007) but focused on the brainstem (Singh et al., 2022)).

### Central Autonomic Network

We studied central ANS dysfunction across an *a priori* autonomic network comprised of eight regions-of-interest (ROI), grouped into four domains:

1. **Medullary** (N=2): Viscero-sensory motor nuclei complex (VSM) and rostro-ventrolateral medulla (RVLM).
2. **Relay** (N=2): Medial parabrachial nuclei (MPB) and periaqueductal grey (PAG).
3. **Hypothalamus** (N=1).
4. **Cortical** (N=3): Central medial nucleus of the amygdala (CeA), anterior insular cortex (AIC), and subgenual anterior cingulate cortex (sgACC).

The VSM was chosen primarily to include the nucleus of the solitary tract, the principal nuclei receiving and relaying afferent autonomic information (‘Basics of Autonomic Nervous System Function’, 2019), but also includes the neighbouring dorsal motor of the Vagus (efferent track) which is indissociable at this resolution. The RVLM was included due to its role in sympathetic up-regulation of blood pressure (Guyenet, 2006). The MPB and PAG were included as they are both major integrator and relay centres for autonomic responses (‘Central Control of Autonomic Function and Involvement in Neurodegenerative Disorders’, 2013). The hypothalamus due to the role of various subnucleus in autonomic control, for example, both the paraventricular and dorsomedial nuclei output to the adrenocortical axis and regulate cerebral blood flow and cardiovascular functions, amongst others (‘Central Control of Autonomic Function and Involvement in Neurodegenerative Disorders’, 2013). The CeA was included as it is the sub-division of the amygdala most connected to the hypothalamus and brainstem, and implicated in autonomic processing (Avecillas-Chasin et al., 2023). The AIC was included due to its role in the sympathetic and parasympathetic regulation (De Morree et al., 2016). Finally, while recognising the complexity of the ACC’s cytoarchitecture and function, we took the sgACC over the perigenual subdivision, given its more recognised role in interoception (Myers et al., 2025) and autonomics (Rudebeck et al., 2014), however, together with the AIC, these two regions should be seen as the “top” end of this ANS network.

Given that this study was conducted at 3T, we averaged bilateral ROIs to boost signal-to-noise ratio. We now describe how these regions were defined and, in some cases, approximated. The VSM was based on the Brainstem Navigator 1.0 (The Brainstem Imaging Laboratory, MIT), but given the resolution of the present work, was approximated by a sphere equidistant from the bilateral locations identified in the Brainstem Navigator. The RVLM was approximated with two spheres, whose locations were approximated from ROIs of the WIKIBrainStem (Lechanoine et al., 2021), an 11.7T study of the brainstem. The MPB was based on the Brainstem Navigator delineation but was again approximated due to resolution and comprised two spheres centred at the respective centre coordinates of the Brainstem Navigator delineations. The PAG was taken directly from the Brainstem Navigator. The CeA was taken from a connectivity-based probabilistic tractography parcellation of the amygdala that distinguished the centromedial, basal and lateral nuclei (Avecillas-Chasin et al., 2023). The AIC was taken from a probabilistic atlas of the insula (Faillenot et al., 2017) (anterior inferior part of the insula, including the apex). The sgACC was taken directly from the Automated Anatomical Labelling Atlas (Rolls et al., 2020).

### Spectral Dynamic Causal Modelling

#### Individual Level Models

For each participant, we recovered the ROIs’ timeseries by masking them with a white and grey matter mask and computing their first principal component. We then used these to estimate effectivity connectivity via the spectral dynamic causal modelling (DCM) implementation in SPM12 (see: https://www.fil.ion.ucl.ac.uk/spm/software/spm12/). At this level participants have a generative model specified with fully connected ROIs. Estimating these models’ parameters is done using variational Laplace, a method which approximates model evidence via free energy and uses it to compare models in an attempt to achieve a trade-off between and accuracy of fit and model complexity (K. Friston et al., 2007; Zeidman, Jafarian, Corbin, et al., 2019). We used the default settings in the SPM12 version of SPM. For further technical details on spectral DCM, see *Friston et al.* (K. J. Friston et al., 2014).

#### Group-level Analyses

Having obtained the optimal parameters for each individual model, we then used parametric empirical Bayes (K. J. Friston et al., 2016) to model variability at the group-level. Parametric empirical Bayes is a hierarchical framework where dependencies from the group-level inform the individual-level and where hypotheses about groups effect can be tested. Importantly, it let us define both the baseline architecture of the autonomic network, specifically characterising the strength and direction of connections that are common across individuals, as well as how connectivity strength in that architecture changes due differences across individuals.

Using this framework, we started by confirming our autonomic network and connectivity hypotheses were valid across both PD and HC, while accounting for age. Then, we separately tested PD and HC for baseline commonalities in autonomic connectivity (group mean), and differences related to steeper drop in systolic blood pressure during the standing task (laying blood pressure minus standing blood pressure at the end of the 3-minutes), while correcting for age and use of blood pressure regulating medication (binary yes or no use). These comparisons represent fundamental *a priori* questions. Default parameters from SPM12 were used and further details on parametric empirical Bayes may be found in *Friston, et al. 2016* (K. J. Friston et al., 2016).

At this stage hypotheses about the architecture of the baseline autonomic network and differences underpinning any covariate can be tested by “turning off” connections of the full individual models in a process known as Bayesian model reduction. We did this in two ways. First, we established factors that explored specific hypotheses about group and directionality of the effects:

1. **Intra-connectivity** between groups of regions, which reflects changes in dynamics between the autonomic network ROIs within each domain (i.e. medulla, relay, cortical).
2. **OUT-connectivity** from specific groups of regions, which reflects the influence of those autonomic on to other areas.
3. **IN-connectivity** into specific groups of regions, which reflects regulation of these groups of autonomic regions.

To operationalise this, we specified groups of models that explored the different factors (Fig. 1A-C), as well as different combinations of the factors (Fig. 1D). Self-connections never change from their intrinsic self-inhibitory assumption. The first factor contained six families, (i) intra-connectivity *on* only for medullary regions, (ii) intra-connectivity *on* only for relay regions, (iii) intra-connectivity *on* only for cortical regions, (iv) intra-connectivity *on* for medullary and relay regions, (v) intra-connectivity *on* for medullary and cortical regions, (vi) intra-connectivity *on* for relay and cortical regions, and (vii) intra-connectivity *on* for all regions. There was no specific family for the hypothalamus given that it is the only region in that group and such a model would be identical to the null model (i.e. only self-connections turned on). The second factor had 16 different families, starting from the null mode, followed by the simplest four families, i.e. OUT-connections from medullary regions, OUT-connections from relay regions, OUT-connections from the hypothalamus, OUT-connections from cortical regions, followed by all possible combinations of these until the full model. The third factor was constructed in the same manner as the previous one, but for IN-connections. All possible combinations with duplicates removed, totalled 1163 models, including null and full models.. Bayesian model comparison was used to compute model evidence for each model, and Bayesian model averaging pooled evidence across models of the same family. The latter implied that obtaining estimates of connection parameters on pooled models, was done by averaging each parameter while weighting it by its model’s posterior probability. We further applied a 95% posterior probability on the parameters being present versus absent in the model. Doing so to jointly explain the baseline across individuals and the differences meant exploring a 1163x1163 model-space but guaranteed a conservative approach in which the obtained posterior probability was in relation to the entire model space (as compared to approaches that split explanatory hypotheses). The second way, which we used to validate the previous results was a hypothesis-free way in which all parameters are turned *on* or *off* to generate all possible combinations, and model evidence is then compared to determine which parameters offer explanatory value. In this case, the parameters from the best 256 models were averaged and weighted by their model posterior, maintaining the threshold of 95% probability of being present on the parameters.

**Figure 1.**
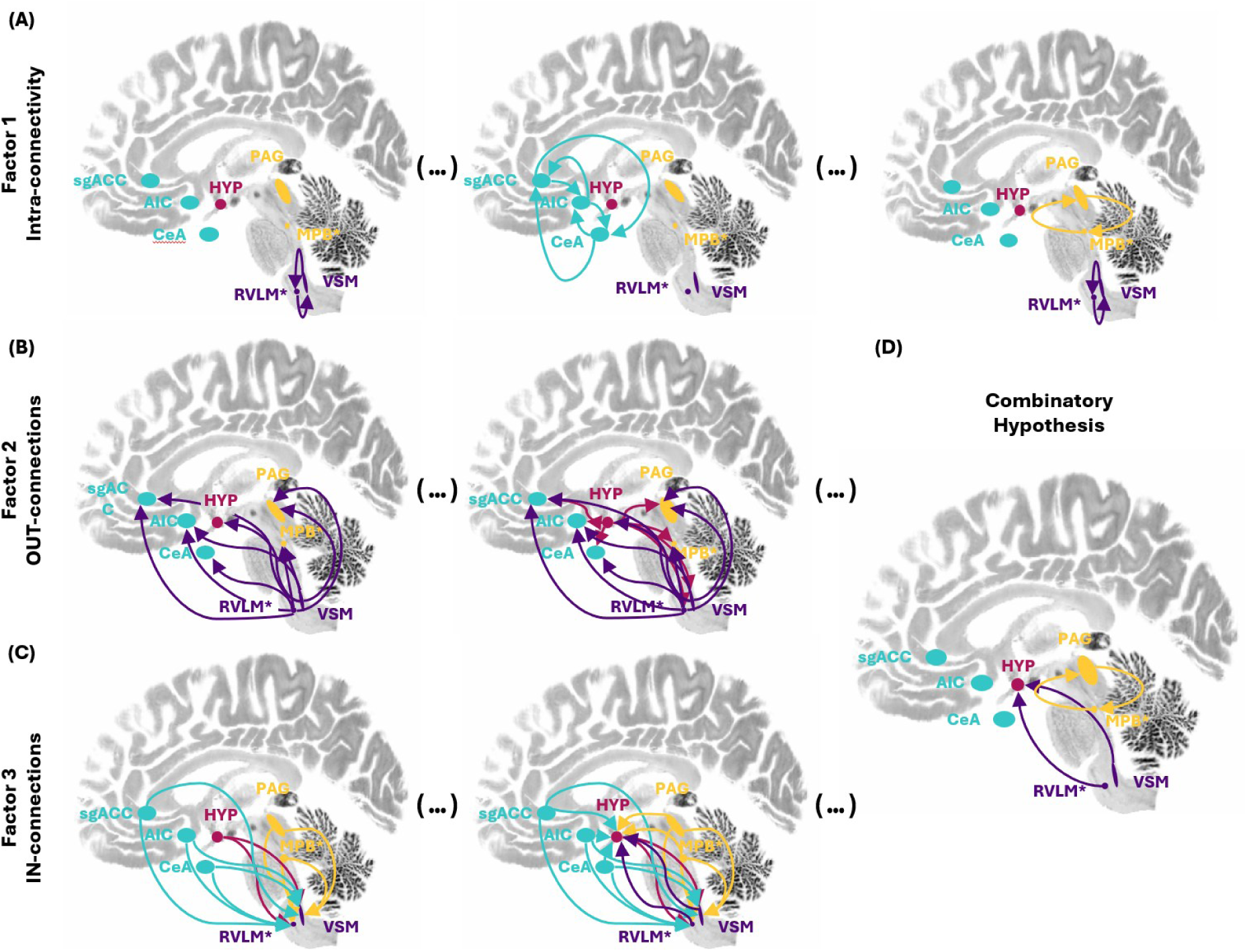
Hypotheses testing for the central ANS network. Three factors tested reduced model hypotheses about the ANS network. All included inherently self-inhibitory connections not represented here. (A) Factor 1 assessed intra-connectivity within types of regions. (B) Factor 2 assessed the role of all connection going out of a specific region type for both individual regions types and their combinations. (C) Factor 3 assessed the role of all connections going into a specific region type, both for individual regions types and combinations. (D) Merging these factors allowed to test specific combinatory hypotheses such as the represented example which is a merger of intra-connectivity within relay regions, out-connections from medullary regions and in-connections to the hypothalamus.

## Data availability

Imaging and clinical data used in this study will be shared upon reasonable request to the corresponding author. All data and statistics generated from this study are presented in the manuscript.

## Results

### Participants

The 81 individuals with PD who participated in the study were on average 59.55 years-old (SD: ±9.38) and had been diagnosed with PD within the two years prior to participating in the study (1.20 ± 0.67). The HC group was on average 61.45 years old (SD: ±8.84). Both groups had equivalent cognitive MoCA scores (Table 1) and had relatively low autonomic dysfunction scores on the SCOPA-AUT (Table 1), albeit slightly higher for the PD group (*P = 0.0145*). 19 out of the 81 individuals with PD and 14 out of the 65 HC used blood pressure medication.

**Table 1.** shows the full patient demographics.

| Metric | PD |  | HC |  | *p-value* |
| --- | --- | --- | --- | --- | --- |
| | Mean ± SD | Range | Mean ± SD | Range | |
| Age | 59.55 ± 9.38 | [32.87; 73.90] | 61.45 ± 8.84 | [35.23; 75.74] | 0.2143 |
| Disease duration | 1.2 ± 0.67 | [0; 4.59] | - | - | - |
| UPDRS-III | 18.89 ± 8.68 | [5; 41] | 1.12 ± 1.55 | [0; 7] | < 0.0001 |
| H&Y | 1.27 ± 0.5 | [0; 2] | 0 | 0 | < 0.0001 |
| SCOPA-AUT | 10.96 ± 6.33 | [0; 31] | 7.13 ± 6.14 | [0; 22] | 0.0145 |
| SCOPA-AUT (CVS) | 0.65 ± 0.80 | [0; 3] | 0.23 ± 0.43 | [0; 1] | 0.0197 |
| MoCA | 26.96 ± 2.35 | [19; 30] | 27.04 ± 1.99 | [22; 30] | 0.8199 |
| Blood pressure medication | 19 (23.46%) | - | 14 (21.54%) | - | - |

### 3-minute standing task

To assess nOH, study participants performed a short standing task in which they laid supine on a medical bed for 5 minutes, with blood pressure and heart rate being measured at the end, and then stood up-straight for 3 minutes, with blood pressure being measured every minute. For individuals with PD, the systolic blood pressure at the third minute standing up showed a change of -5.46 mmHg (SD: ±10.11; range: -35 to +15) as compared to the one laying down, taken just before standing up (Fig. 2). The HC group showed a change of -4.60 mmHg (SD: ±11.82; range: -35 to +30), which was not statistically different in mean (*P = 0.6377*) nor variance (*P = 0.1840*) from the one experienced by the PD group.

**Figure 2.**
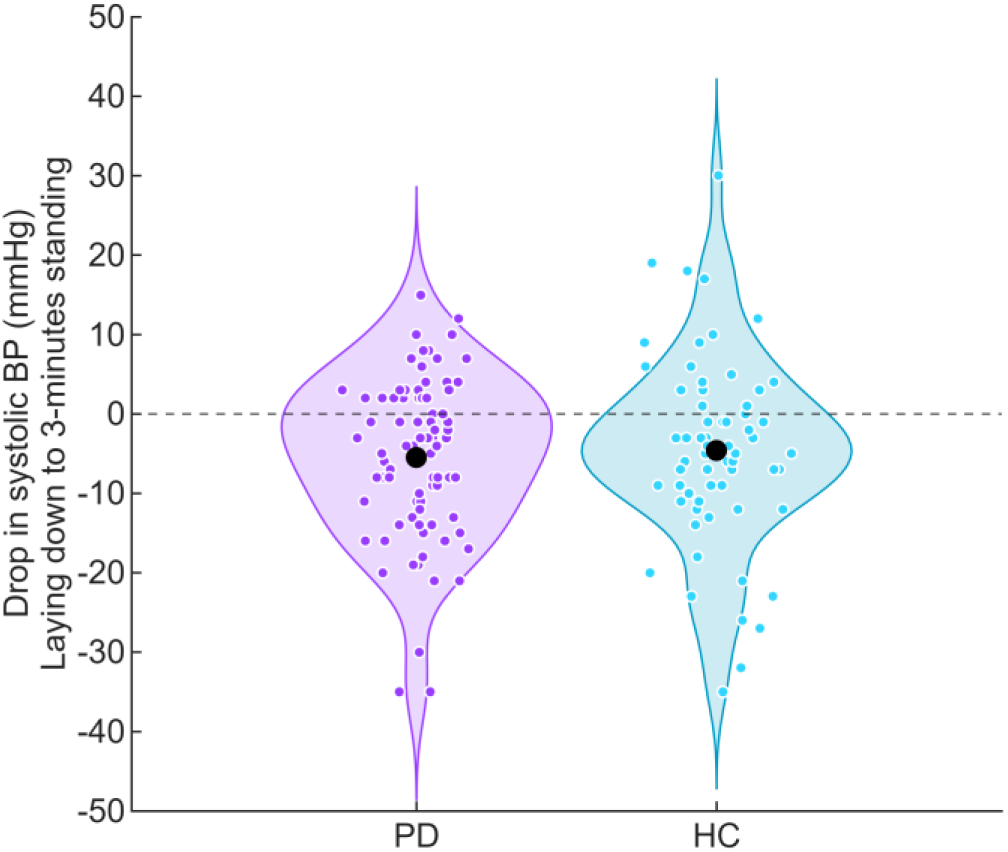
Blood pressure outcomes from the standing task. The violin plots show the distributions of the change in systolic blood pressure from the end of 5-minutes laying down to the end of 3-minutes standing, in purple for PD and teal for HC. Single points represent individual participants. Individuals with PD experienced an average decrease in systolic blood pressure of -5.46 mmHg (black dot in the purple distribution) and the healthy participants experienced an average decrease of -4.60 mmHg. This was not significantly different between the two populations.

### Accuracy of the individual dynamic causal models

Across all patients and healthy participants, the individual-level DCM estimations had an explained variance of 89.14 ± 2.49% (range: 80.91% to 95.30%) when fitted to the observed spectral data, which demonstrated a good-fit between estimated and actual cross-spectra.

### Evidence supporting a common autonomic network

As a preliminary step we validated that across PD and HC there was significant evidence supporting the central autonomic network under investigation. Over the possible 1163 models for a baseline architecture common to PD and HC, model #1161 showed a posterior probability of 98.71%, demonstrating we could identify well a common autonomic network across PD and HC. This network was characterized by intra-connectivity within all groups, OUT-connectivity from the medulla, hypothalamus and cortex, and IN-connectivity into all groups. Analysing blood pressure effects had to be done separately per group due to model complexity.

### Evidence for group-level models in PD

To examine the autonomic network’s baseline architecture across individuals with PD, as well as differences related to changes in systolic blood pressure during the standing task, we used parametric empirical Bayes to specify group-level effects (baseline and differences) and Bayesian model comparison to assess evidence for various connectivity hypotheses. Specifically, we hypothesized effective connectivity across the autonomic network’s four groups of regions, i.e. medullary (VSM and RVLM), relay (MPB and PAG), hypothalamus, and cortical (CeA, AIC, sgACC); would change with regards to intra-connectivity, OUT-connectivity, and IN-connectivity.

Across the 1163-by-1163 model space built to explain baseline and differences, the highest joint posterior was 40.99% and belonged to models #1114 for the baseline and #240 for the differences (Fig. 3). The support for these models to explain only baseline or only differences (i.e. marginal posteriors) was 68.04% and 60.30%, respectively (Fig. 3). Both joint and marginal posteriors were considerably high in the context of *evidence dilution* brought by the high number of models (random probability: ∼8x10^-7^ per *joint* models, ∼8x10^-4^ per *marginal* model), however, not reaching 90% meant no model provided a *unique* solution for the data across this hypotheses space.

**Figure 3.**
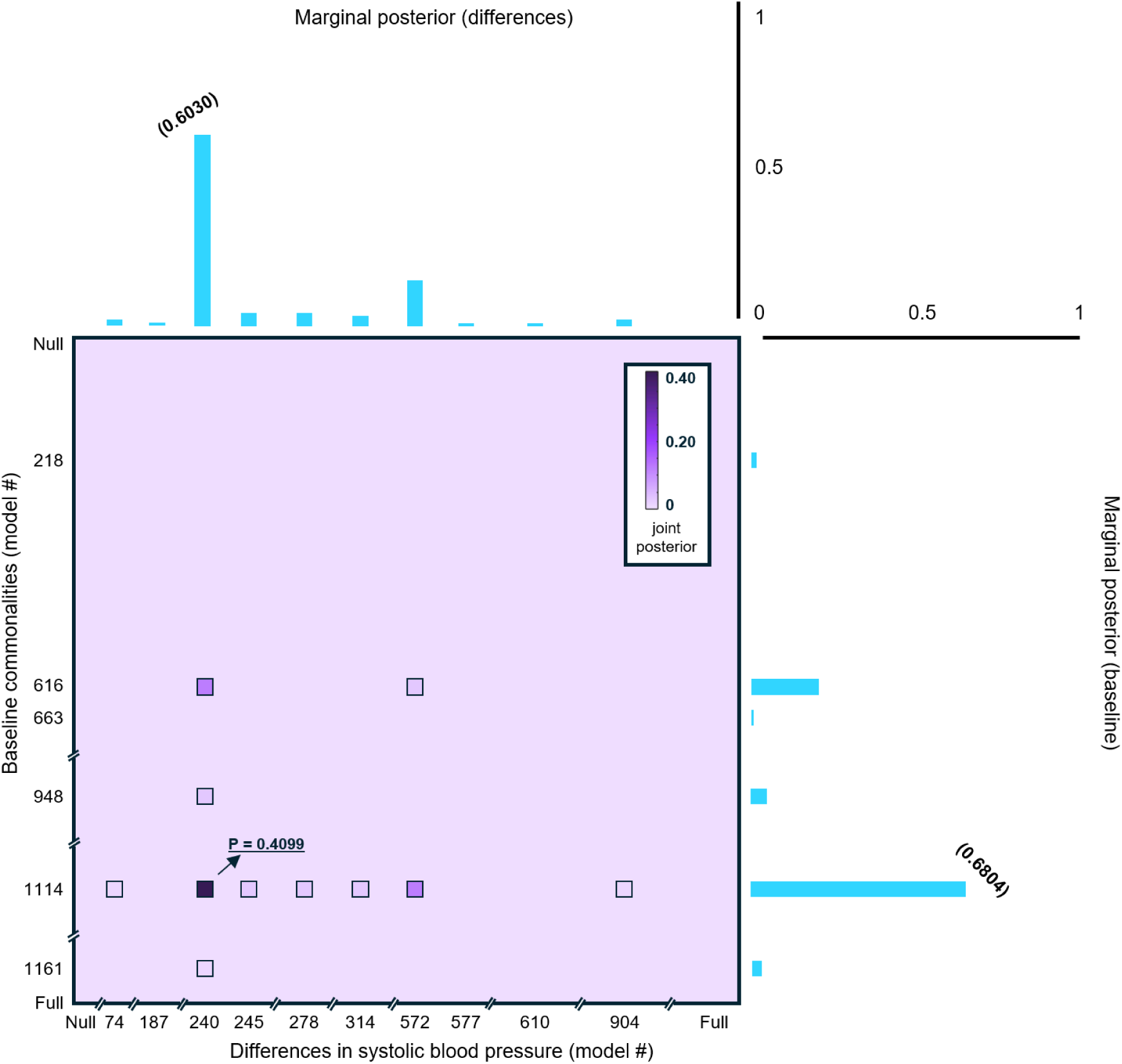
Posterior evidence for all combinations of models explaining commonalities and differences in connectivity due to drops in blood-pressure in PD. Merging our connectivity hypotheses (intra, OUT, IN) for the central ANS network, led to 1163 possible arrangements of connectivity. These models were used to explain both the commonalities across individuals with PD and the differences associated with drops in blood pressure during the standing task. The large square represents the entire model space and their associated joint-posteriors, which for most models is close to zero (very large model-spaces lead to evidence dissolution). Smaller squares within the space represent models which reached at least a 1% joint-posterior. The combination of models #1114 for the baseline and #240 for differences reached a joint-posterior of 40.99%, which is very high considering the model-space. Blue bars on the right represent the evidence for baseline models alone, which is achieved by summing (i.e. marginalising) over the differences’ models (e.g. for baseline model #1161 the marginal-posterior is obtained by summing over all difference models). Top blue bars represent the same but for differences.

To better understand the baseline architecture and differences in connectivity, we assessed model evidence across our predefined hypotheses for intra, OUT- and IN-connectivity. With respect to baseline, the highest marginal family-posteriors were observed for intra-connectivity within all groups of regions (marginal family posterior: 70.95%; Fig. 4A), OUT-connectivity from the medulla, hypothalamus and cortex (marginal family posterior: 99.98%; Fig. 4B), and IN-connectivity into the medulla, relay and cortical regions (marginal family posterior: 95.92%; Fig. 4C). The conjunction of these three hypotheses corresponded to model #1114 for the baseline architecture, but the hypotheses for OUT- and IN-connectivity were the ones garnering the strongest evidence to explain the baseline architecture, with the former being highest. With respect to differences in connectivity, the highest marginal family-posteriors corresponded to intra-connectivity within relay regions (marginal family posterior: 75.99%; Fig. 4A), OUT-connectivity only from the medulla and hypothalamus (marginal family posterior: 91.42%; Fig. 4B), and IN-connectivity into relay and cortical regions for the differences (marginal family posterior: 85.99%; Fig. 4C). The conjunction of these three predefined hypotheses corresponded to model #240 for the differences in connectivity, but only OUT-connectivity garnered strong evidence to explain the differences. As such, our final joint explanatory model was defined by the strongest explanatory families for baseline and differences, which for both cases was OUT-connectivity from the medulla and hypothalamus, as well as cortex for the baseline.

**Figure 4.**
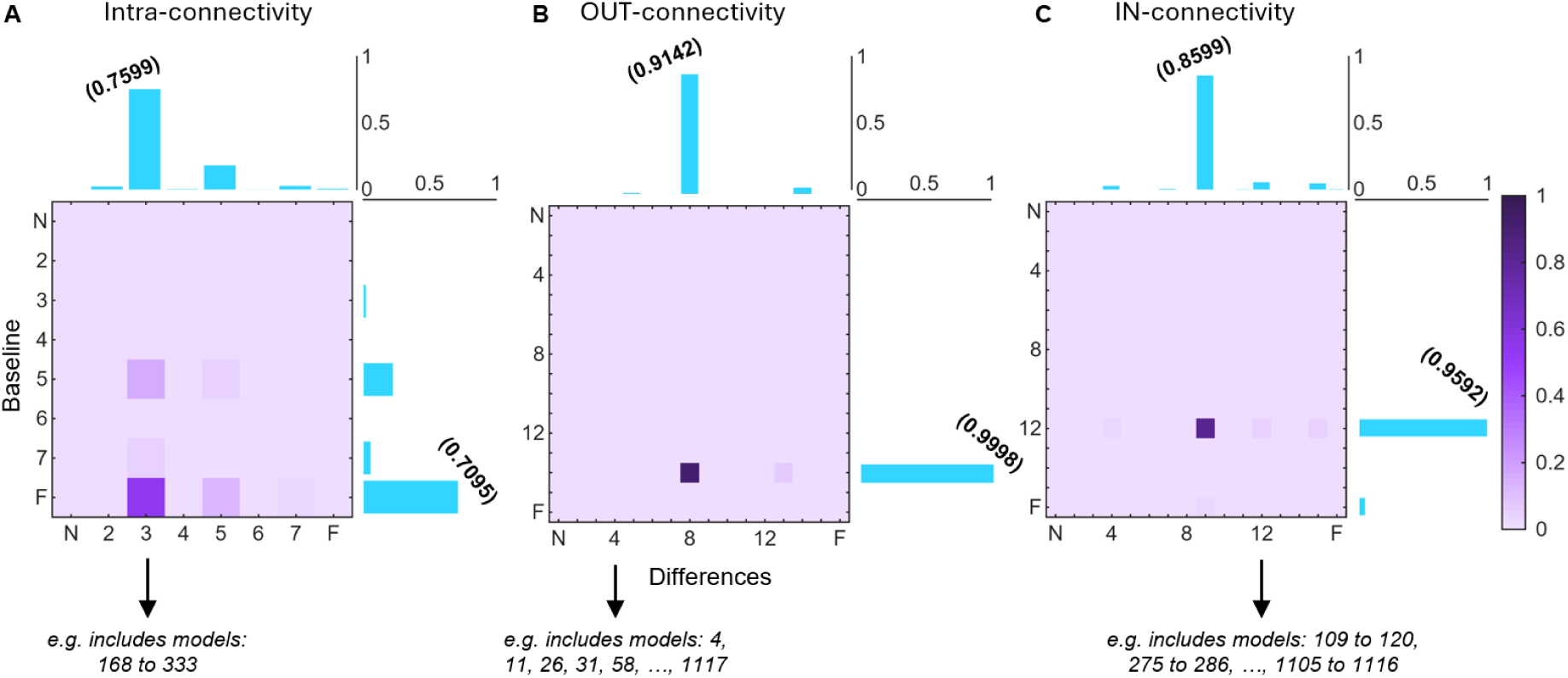
Posterior evidence for families of models in PD. All models used to explain differences in connectivity came from a specific family of the hypothesised factors (see Figure 1). Here, instead of showing joint-posterior for each specific model, evidence is pooled across models of the same families to give interpretability to the observed changes. For example, model #240 for the differences is a specific variation of the factors which combines the 3^rd^ family of intra-connectivity (that is intra-connectivity in relay regions only), with the 8^th^ family of OUT-connectivity (that is OUT-connectivity from the medulla and hypothalamus) and 9^th^ family of IN-connectivity (that is IN-connectivity into relay nuclei and cortex). Pooling across families can reveal that model evidence is in fact not as spread out as it might seem. For example, for intra-connectivity (A) marginal family posteriors show 75.99% support for the 3^rd^ family for the differences, and 70.95% support for the full model for the baseline. For OUT-connectivity (B) the 8^th^ family has a 91.42% family marginal posterior for the differences and 99.98% for the 14^th^ family (OUT-connectivity from the medulla, hypothalamus and cortex). Lastly, IN-connectivity shows 85.99% family marginal posterior for the 9^th^ family for the differences, and 95.92% for the 12^th^ family for the baseline (IN-connectivity into the medulla, relay and cortical regions).

### Baseline network architecture in individuals with PD

The structure of the baseline network architecture as defined by the final joint explanatory models, was obtained by performing a weighted average of the connection parameters by each model’s posterior (Bayesian model averaging). Figure 5A shows the baseline architecture common to individuals afflicted by PD, with all parameters satisfying a 95% probability for being present versus absent from the model. The autonomic network was overall marked by an inhibitory pattern (as in activation in one area leads to deactivation in another – the underlying neurochemical composition is not inferred), and here we report the ones with above 5% change. The RVLM presented inhibitory input into most of areas, including AIC (-0.0796), CeA (-0.1118), PAG (-0.0986), and sgACC (-0.0784). The VSM also showed overall inhibitory output, particularly to the MPB (-0.0617) and PAG (-0.0736), and otherwise presented weak self-inhibition (-0.0768). The hypothalamus showed a clear distinction between inhibitory connectivity to the brainstem, such as to the RVLM (-0.0787) and VSM (-0.0857), and excitatory projections to the cortex, such as to the CeA (+0.1129) and sgACC (+0.0914). Additionally, the hypothalamus showed markedly decreased self-inhibition at baseline (-0.1278). Cortical regions mostly exhibited concordant inhibition to medullary and relay areas, e.g. AIC to MPB: -0.0715; CeA to VSM: -0.0736; sgACC to all brainstem regions < -0.07. Also in the cortex, the AIC showed decreased self-inhibition (-0.1146). Finally, within relay regions the PAG showed strong inhibition towards the MPB (-0.1711) and the MPB showed decreased self-inhibition (-0.1707) despite various inhibitory input. Lastly, to validate this hypotheses-based analysis, we ran an automated parameter search, which supported the estimations of the neuronal parameters obtained here.

**Figure 5.**
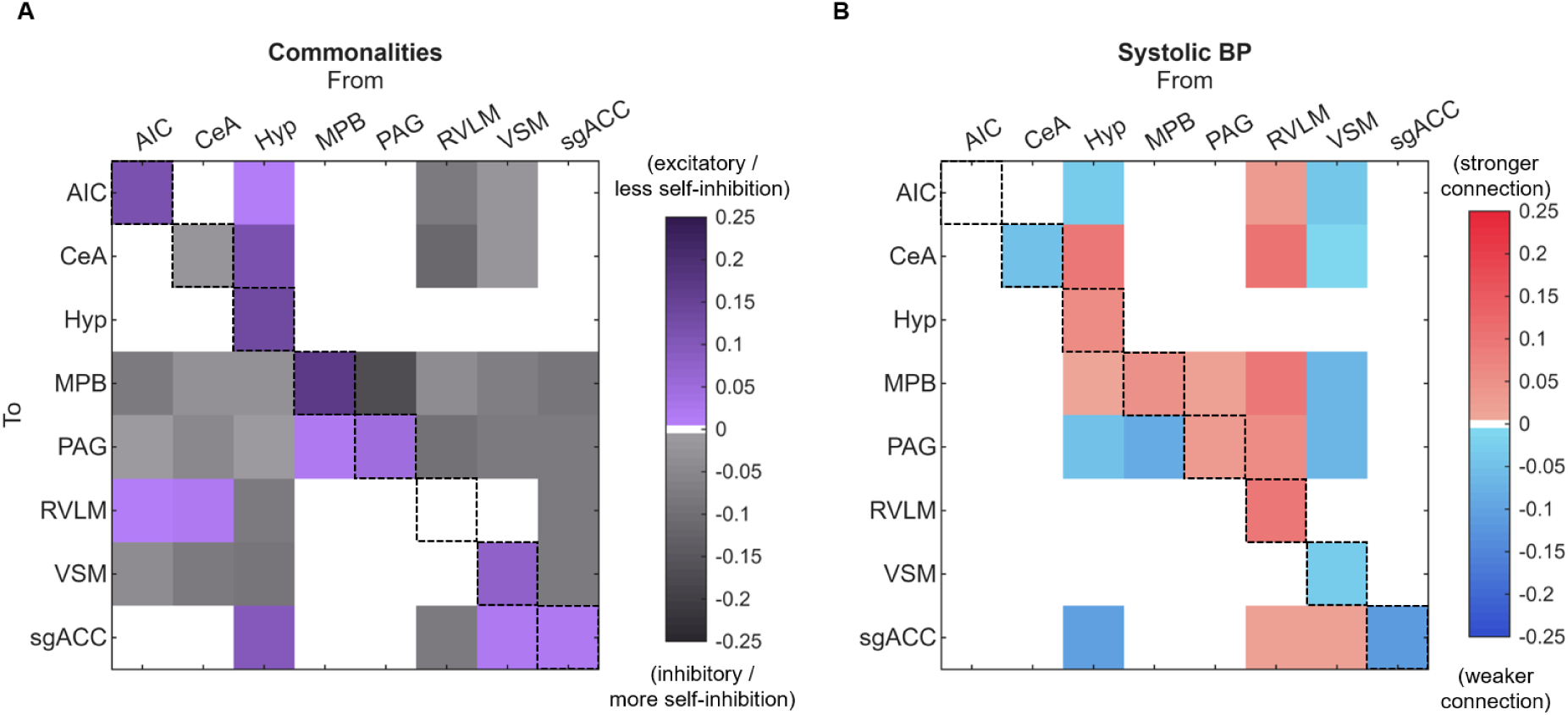
Estimated neuronal connectivity parameters in PD. Bayesian model averaging across the families of models with highest posterior revealed (A) the baseline architecture of the central autonomic network, as well as (B) the changes associated with drops in blood-pressure during the standing task. **(A)** The baseline architecture is marked by an inhibitory pattern from the medulla to relay and cortical regions, and reciprocal inhibition from the cortex to medullary and relay regions. The hypothalamus shows a clear distinction between inhibition to the brainstem, and excitation to the cortex. It should be noted that excitation in this context means that higher activity in one region leads to higher activity in another, whilst inhibition means that higher activity in one region leads to lower activity in another. The nature of the underlying synaptic transmission is not inferred and there may be intermediate populations. **(B)** Alterations in connectivity at baseline associated with drops in blood pressure, identify a pattern of increased inhibitory connections departing from the RVLM. In addition, there is a weakening of inhibition from the VSM and overall increase in inhibition within the relay nuclei that may decrease the activity of this group. ***<u>For DCM experts:</u> We are aware that in classical DCM output, negative diagonal elements mean decreased self-inhibition and positive mean increased self-inhibition. We flipped these values in figure A to improve interpretation. Hence, for the purposes of this matrix only, positive values represent weaker self-inhibition; negative values represent stronger self-inhibition*.**

### Connectivity differences with drops in blood pressure in PD

Alterations in connectivity related to changes in systolic blood pressure during the standing task were also obtained via Bayesian model averaging on the final joint explanatory models (Figure 5B). Interpreted in relation to the inhibitory/excitatory nature defined by the baseline network architecture, the most prominent connectivity changes were seen from medullary regions and from the hypothalamus (Figure 6A). With respect to medullary changes, these include stronger inhibition from the RVLM to the AIC (+0.0993), MPB (+0.0925), PAG (+0.0584), and to itself (i.e. increased self-inhibition; +0.0968), and decreased inhibition from the VSM to the MPB (-0.0633) and PAG (-0.0652). With respect to the hypothalamus, we observed increased excitatory input to the CeA (+0.0928), increased self-inhibition (+0.0553), decreased inhibitory input to the PAG (-0.0460), and decreased excitatory input to the sgACC (-0.1038). Other notable changes included decreased self-inhibition of the CeA (-0.0449), increased self-inhibition of the MPB (+0.0485), decreased excitatory input from the MPB to the PAG (-0.0819), and decreased self-inhibition of the sgACC (-0.1116). As before, we validated these results using a hypotheses-free automated parameter search, which confirmed the main estimated parameters by the family-based hypotheses.

**Figure 6.**
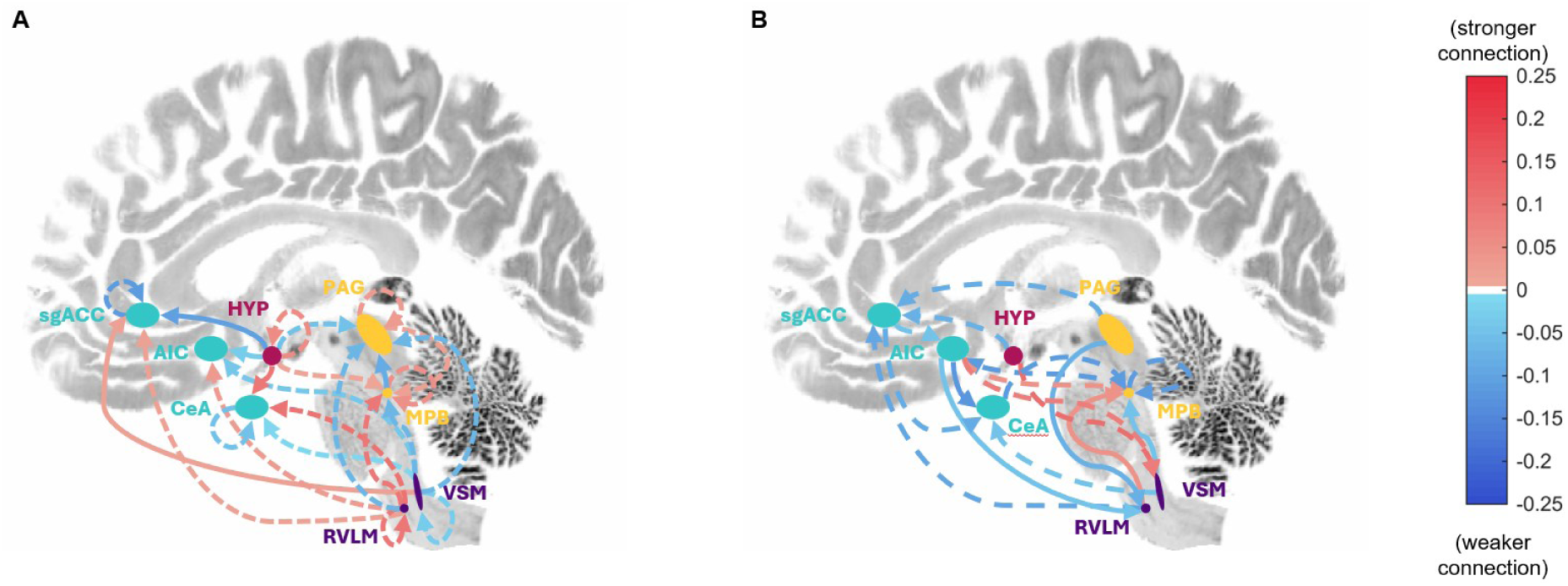
Representation of neuronal changes due to drops in blood pressure. We overlayed the changes in connectivity associated with higher drops in blood pressure during the standing task, with the identified nature of the connection. Solid lines represent excitatory connectivity while dashed lines represent inhibitory connectivity, and red lines indicate connection that got stronger in associated with the blood pressure drop whereas blue lines indicate connections that got weaker. **(A) PD:** In this format the bottom-up pattern of stronger RVLM inhibitory influence on other regions becomes noticeable and so does the weaking on inhibition from the VSM. The hypothalamus shows weakening inhibitory connections to the brainstem, and excitatory to the cortex, and interestingly excitation decreases for the AIC and sgACC, but increases to the CeA. Given the increased inhibition on the CeA from the RVLM, this may represent a compensatory mechanisms from the hypothalamus. **(B) HC:** We observe a weakened pattern of connectivity with strong drops in blood pressure, however, there does not seem to be any specificity to groups of brain regions as was noticeable in PD.

### Evidence for group-level models in HC

The same analysis for HC showed that across the model space, the highest joint posterior was 25.28% and belonged to the full model #1163 for the commonalities and model #831 for the differences. Family hypotheses testing, with respect to the baseline, supported full intra-connectivity across all regions (marginal family posterior: 91.75%), full OUT-connectivity from all regions (marginal family posterior: ∼100%), and full IN-connectivity into all regions (marginal family posterior: ∼100%). With respect to the differences, it supported intra-connectivity within the medulla and cortex (marginal family posterior: 37.98%), OUT-connectivity from all regions (marginal family posterior: 87.39%), and IN-connectivity into all regions (marginal family posterior: 81.43%). Hence, the joint explanatory model for HC was reduced to the combination of the full OUT and IN connectivity from all regions for the baseline, as well as OUT-connectivity from all regions for the differences.

### Baseline network architecture in healthy individuals

Bayesian model averaging across the final joint explanatory model revealed that, similarly to PD, the baseline network architecture in HC was marked by an overall inhibitory pattern. Figure 7A shows the connectivity values for a 95% probability of being present versus absent. Here, we report the ones above 5% change. In the medulla: the RVLM presented inhibitory input to the AIC (-0.1280), CeA (-0.1692), hypothalamus (-0.1522), PAG (-0.1534) and sgACC (-0.0834), excitatory input to the VSM (+0.1732) and overall less self-inhibition (-0.0852); and the VSM presented inhibitory input to the PAG (-0.0537) and excitatory input to the sgACC (+0.0597). For relay nuclei: the MPB showed inhibitory input to the AIC (-0.0510) and hypothalamus (-0.0806) and much decreased self-inhibition (-0.2174); and the PAG showed inhibitory input to the AIC (-0.1145), excitatory input to the VSM (+0.1053), and weaker self-inhibition (-0.1777). The hypothalamus show inhibitory input to the VSM (-0.2263), excitatory to the MPB (+0.0603) and PAG (+0.0676). Finally, for cortical regions: the AIC showed inhibitory input to the VSM (-0.0537) and excitatory to the sgACC (+0.1277); the CeA showed inhibitory input to the MPB (-0.0741) and PAG (-0.0742); and the sgACC showed inhibitory input to the AIC (-0.1402), hypothalamus (-0.0769), MPB (-0.0829), PAG (-0.0898), RVLM (-0.1077), and VSM (-0.1126). An hypotheses-free automated parameter search, supported the estimations obtained here for the neuronal parameters.

**Figure 7.**
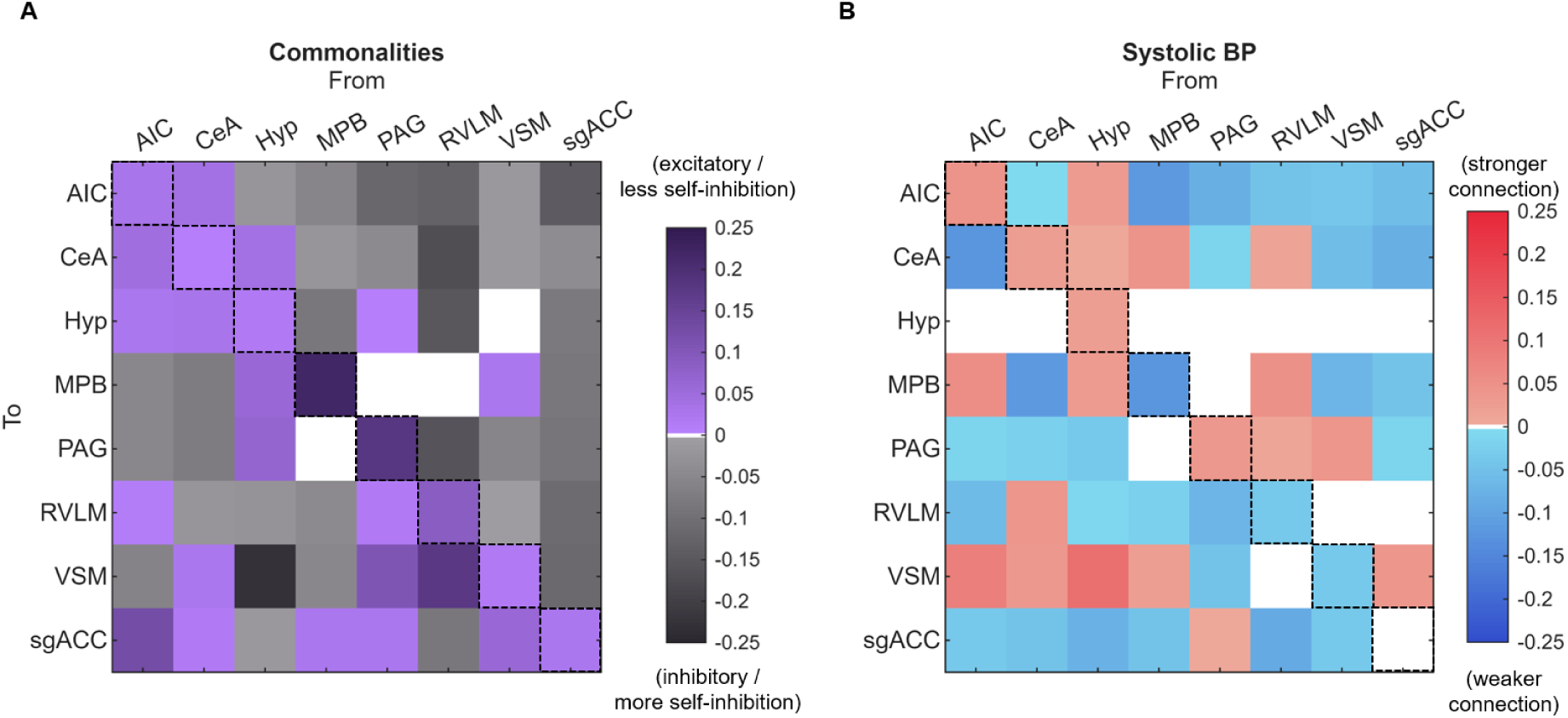
Estimated neuronal connectivity parameters in HC. Bayesian model averaging across the families of models with highest posterior revealed (A) the baseline architecture of the central autonomic network, as well as (B) the changes associated with drops in blood-pressure during the standing task. **(A)** The baseline architecture observed in HC is very similar to that of PD, mostly marked by inhibition in between the same regions, but does include more excitatory intra-connectivity in the cortex. **(B)** Alterations in connectivity at baseline associated with drops in blood pressure, reveal a much more distributed and unspecific pattern in HC as compared to PD, which may be related to the specificity of changes in this population. However, overall and especially when considering the strongest changes, the pattern may be revealing of an overall weakening of connections regardless of the excitatory or inhibitory nature. ***<u>For DCM experts:</u> We are aware that in classical DCM output, negative diagonal elements mean decreased self-inhibition and positive mean increased self-inhibition. We flipped these values in figure A to improve interpretation. Hence, for the purposes of this matrix only, positive values represent weaker self-inhibition; negative values represent stronger self-inhibition*.**

### Connectivity differences with drops in blood pressure in HC

Again, Bayesian model averaging on the final joint explanatory models revealed changes in central autonomic connectivity related to more pronounced drops in blood-pressure during the standing task (Figure 7B, with 95% threshold of being present versus absent). Interpreted in relation to the inhibitory/excitatory nature defined by the baseline network architecture, these connectivity changes relatively widespread and non-specific (Figure 6B). With respect to medullary changes (only above 5% change reported): the RVLM increased excitatory input to the MPB (+0.0500) and decreased inhibitory input to the sgACC (-0.0868); and the VSM decreased inhibitory input to the CeA (-0.0515) and decreased excitatory input to the MPB (-0.0669). For relay nuclei: the MPB decreased inhibitory input to the AIC (-0.1160) and decreased self-inhibition (-0.1193); and the PAG decreased inhibitory input to the sgACC (-0.0773) and decreased excitatory input to the RVLM (-0.0701). The hypothalamus increased inhibitory input to the VSM (+0.1120) and decreased inhibitory input to the sgACC (-0.0745). Finally, for cortical regions: the AIC decreased excitatory input to the CeA (-0.1234) and to the RVLM (-0.0578), increased inhibitory input to the MPB (+0.0544) and to the VSM (+0.0812); the CeA decreased inhibitory input to the MPB (-0.1178); and the sgACC decreased inhibitory input to both the AIC (-0.0553) and CeA (-0.0801).

## Discussion

We used generative modelling to estimate effective connectivity across a central autonomic network in individuals with early PD and aged-matched controls, and then deployed a comprehensive Bayesian methodology to test how that connectivity changed in relation to drops in blood-pressure during an orthostatic challenge. From these analyses, two main findings emerge. First, the baseline autonomic network in PD and HC is predominantly inhibitory, with the RVLM exerting widespread inhibitory influence on relay and cortical regions, and cortical areas as well as the hypothalamus in turn inhibiting the brainstem. Second, the magnitude of orthostatic blood-pressure drops in PD, was specifically related to increased inhibition from medullary regions and altered hypothalamic output to both cortical and brainstem targets. This was distinct from blood-pressure drops in HC, which showed non-specific and distributed changes, that were not dominated by the bottom-up medullary influence seen in PD.

To our knowledge, these results provide the first *in-vivo* evidence for central autonomic functional reorganisation, in early-PD, consistent with impaired sympathoexcitatory capacity. Such reorganisation at rest may predispose those with PD to dampened central reactivity to orthostatism, which actively contributes to steeper drops in blood-pressure when standing and ultimately to nOH.

### Predominant inhibition across central autonomic regions

The baseline network architecture common to individuals with PD, was predominantly characterised by inhibitory connectivity. This was most apparent in RVLM inhibitory influence on relay nuclei and cortical regions, and in cortical regions that reciprocated inhibitory influence on brainstem structures. Importantly, a similar inhibitory pattern was observed for HC, suggesting this primarily inhibitory architecture may be an organisation principle of central autonomic regions, rather than a pathological feature of PD.

While there are limited data to validate these *in-vivo* findings, they are consistent with known physiology of autonomic blood-pressure regulation. During rest, baroreflex maintains arterial pressure through inhibitory control of the RVLM that is mediated by GABAergic neurons of the caudal ventrolateral medulla (Dampney et al., 2002). Given that the RVLM is the sympathoexcitatory nucleus responsible for tonic activation of preganglionic sympathetic neurons controlling cardiac output and vascular resistance (Guyenet, 2006), this critical inhibition from the caudal ventrolateral medulla prevents unopposed sympathetic drive. Importantly, the latter neurons do more than just relaying signals from baroreceptor-sensitive neurons from the NST, and have an intrinsic inhibitory baseline activity towards the RVLM (‘Central Control of Autonomic Function and Involvement in Neurodegenerative Disorders’, 2013; Guyenet, 2006). Hence, the predominantly net suppressive influence exerted by the RVLM on other parts of the network in resting state, may be broadly consistent with this tonic inhibitory control.

Reciprocal cortical inhibition of brainstem structures may also be consistent with top-down autonomic regulation. The AIC modulates both cardiovascular sympathetic and parasympathetic output (De Morree et al., 2016; Nagai et al., 2010; ‘The Insular Cortex’, 2012), and the subgenual ACC is implicated in interoception and autonomic modulation (Seamans & Floresco, 2022). Their inhibitory influence on brainstem nuclei at rest, may reflect tonic cortical gating of autonomic outflow that constrains brainstem autonomic responses in the absence of physiological challenge, the opposite of their function when maintaining arousal (Rudebeck et al., 2014).

Another notable feature of the baseline architecture was a clear distinction between hypothalamic inhibitory projections to the brainstem and hypothalamic excitatory projections to cortical regions. We hypothesise that this dual-pattern reflects the role of hypothalamic nuclei, such as paraventricular and dorsomedial nuclei, in integrating descending signals that contribute to autonomic control (Saper, 2002; Watts, 2022), with ascending signals that contribute to interoception and arousal (Berntson & Khalsa, 2021; Watts, 2022). The inhibitory influence of the hypothalamus on medullary regions, at rest, may reflect tonic restraint over the sympathoexcitatory outputs that these nuclei trigger under stress or postural challenge (Dampney et al., 2002). Conversely, excitatory cortical projections may maintain the cortical awareness of visceral states that underpin interoception (Saper, 2002).

### Connectivity changes related to blood-pressure drops in PD: evidence for central sympathoexcitatory failure

Steeper drops in systolic blood-pressure during the standing task were associated with increased self-inhibition of the RVLM and of its inhibitory outputs to the AIC, MPB and PAG. We argue that this pattern is consistent with failure of the central sympathoexcitatory response that maintains blood-pressure and is to our knowledge the first direct *in-vivo* description of such a dysfunction, which may make those with PD more susceptible to drops in blood-pressure.

Under normal physiological conditions, standing-up triggers baroreflex-mediated disinhibition of the RVLM and increases sympathetic outflow to the vasculature and heart, adapting blood-pressure to orthostatic stimuli (‘Central Control of Autonomic Function and Involvement in Neurodegenerative Disorders’, 2013; Guyenet, 2006). In PD, failure to adapt blood-pressure to standing, i.e. nOH, is a typical presentation (Mendoza-Velásquez et al., 2019) tied to deficiencies in two peripheral and two central components (Jain & Goldstein, 2012). First, cardiac noradrenergic denervation, which is universal in PD with nOH (Goldstein & Sharabi, 2019; Lamotte et al., 2020), removes the sympathetic terminals that increase cardiac contractibility and sustain heart-rate acceleration, impairing the heart’s contribution to cardiac output during an orthostatic challenge (Goldstein & Sharabi, 2019). Second, extra-cardiac noradrenergic denervation reduces autonomic fibres (Donadio et al., 2014, 2018) and depletes the postganglionic sympathetic terminals in blood vessels throughout the body (Goldstein, Holmes, et al., 2011), directly compromising the vasoconstriction needed to maintain peripheral vascular resistance when standing (Goldstein, Holmes, et al., 2003). Third, centrally, there is failure of both the parasympathetic and sympathetic arms of the arterial baroreflex arc (Jain & Goldstein, 2012). The cardiovagal limb, which withdraws parasympathetic outflow to the heart accelerating heart-rate in response to falling blood-pressure (Wehrweun & Joyner, 2013), operates through a cholinergic pathway independent of noradrenergic innervation, meaning its failure in PD with nOH (Goldstein, Pechnik, et al., 2003) cannot be explained by peripheral denervation and must involve a central dysfunction (Goldstein, 2003). Finally, the sympathoneural limb, the reflexive increase in sympathetic outflow to the vasculature (Wehrweun & Joyner, 2013), also seems to be affected in PD with nOH (Blaho et al., 2017; Goldstein & Sharabi, 2023) but is harder to disentangle from the peripheral denervation that will either way produce a blunted response. However, lowered muscle sympathetic nerve activity was recently demonstrated in early PD even with preserved peripheral sympathetic nerve fibres and preserved peripheral adrenoreceptor sensitivity (Krämer et al., 2019); definitely strengthening the case for central *in-vivo* dysfunction of the sympathoexcitatory groups that participate in this arc. Post-mortem studies have long demonstrated that medullary effectors are the earliest brain structures afflicted by the ascending progressive nature of α-synuclein pathology (Benarroch, 2004; Braak et al., 2003), but it remained uncharacterised how central autonomic regions became dysfunctional *in-vivo*. Now, we have characterized how this central circuitry becomes, *in-vivo,* marked by stronger inhibitory connectivity from the RVLM, including stronger self-inhibition. We propose that this stronger inhibitory pattern at baseline weakens medullary reactivity to orthostatism leading to higher drops in blood-pressure. Finally, this contrasted with the pattern observed in HC, which saw less self-inhibition of the RVLM, was overall more disengaged (weaker connections regardless of excitatory or inhibitory nature) and was more widespread, i.e. not specific to medullary regions nor the hypothalamus. Presumably, this might have been because in aged-matched HC the potential underlying causes are varied (e.g. natural variation or, considering age, a potential undiagnosed condition) they do not translate into a specific neuronal pattern.

Our findings also identified changes in medullary projections unrelated to sympathetic effectors. The NST, the entry point for the baroreflex arc and predominant relay centre of baroreceptor afferents (‘Central Control of Autonomic Function and Involvement in Neurodegenerative Disorders’, 2013), shows reduced inhibitory influence over relay nuclei and cortex. Lower numbers of NST neurons have been described in animal models of cardiovascular autonomic dysfunction in PD (Falquetto et al., 2017), and it is possible that the pattern we observed reflects impaired transmission of baroreceptor information through the central autonomic network, compromising the integrated autonomic response to an orthostatic challenge. Given that at this resolution we could not dissociate the NST from the dorsal motor nucleus of the Vagus, the first afflicted brainstem nuclei (Benarroch et al., 2006; Braak et al., 2003; Cersosimo & Benarroch, 2008), it is also possible that those changes reflect other pathological changes. However, the directionality observed is more compatible with an afferent receiving and relaying information, than with an efferent nuclei like the dorsal motor of the Vagus.

### Beyond the medulla: compensatory or dissociated hypothalamic responses and constrained relay function

Beyond the medullary changes discussed above, the connectivity alterations associated with steeper blood-pressure drops in PD involved prominent changes in the hypothalamus and relay nuclei that suggest a broader disruption of the integrative middle-tier of the central autonomic network.

The hypothalamus acts as a brainstem-cortex bridge, translating brainstem autonomic states into cortical representations, and channelling cortical and limbic influences to the effector nuclei that control cardiovascular output (Berntson & Khalsa, 2021; Watts, 2022). Here, steeper blood-pressure drops were associated with a specific divergence in hypothalamic output to the cortex. While excitatory drive decreased to the AIC and sgACC, it increased specifically towards the CeA. The CeA is the main autonomic division of the amygdala, with direct projections to brainstem autonomic nuclei (Amunts et al., 2005; Avecillas-Chasin et al., 2023). The up-regulation of this connection may represent a compensatory attempt, already at baseline, to recruit amygdala-brainstem pathways typically engaged during threat and stress, in response to the failing of the primary medullary sympathoexcitatory route. Interestingly, the concurrent decrease in excitation of the sgACC and AIC, may be a contributing factor via dysregulation or visceral interoceptive habituation, to the unawareness of nOH observed in many individuals with PD (Lenka et al., 2024; J. Palma et al., 2015) even with severe blood-pressure drops (Arbogast et al., 2009).

Relay nuclei showed further compromise to the central network integrative capacity. The MPB showed increased self-inhibition, constraining a node that at baseline showed one of the least self-inhibitory patterns – likely related to the physiological convergence of visceral afferent signals requiring high-throughput relay (‘Central Control of Autonomic Function and Involvement in Neurodegenerative Disorders’, 2013). Similarly, the PAG also showed higher self-inhibition potentially compromising its ability to generate autonomic reactions(Deakin & Graeff, 1991; George et al., 2019). This was compounded by increased inhibition of the MPB by the PAG, and a concomitantly weaker excitation of the PAG by the MPB. The net effect is a relay system simultaneously more constrained in its throughput and perhaps less reactive to other nuclei or cortical input, impairing the coordinated multi-level autonomic response required to adapt blood pressure during postural challenges.

### Methodological considerations and limitations

While these results represent the first *in-vivo* characterization of effective connectivity in a central autonomic network in PD, several limitations should be acknowledged. First, fMRI data was acquired at the rest, while blood-pressure measurements were obtained during an orthostatic task outside the scanner. As such, the connectivity patterns associated here with steeper-drops in blood-pressure, reflect trait-level vulnerability rather than real-time central autonomic responses. Second, this study was conducted at 3T fMRI constraining the precision at which small brainstem nuclei could be delineated. This led to the approximation of several nuclei, such as the RVLM, MPB and VSM, and meant that neighboring regions were very much included in the delineated regions-of-interest. Moreover, bilateral averaging of these regions, to boost signal-to-noise ratio, meant losing interpretability of bilateral effects. One solution to this would be via ultra-high-field MRI to improve spatial resolution coupled with longer acquisitions to further boost SNR. Third, effective connectivity as estimated by spectral DCM reflects how one region cause increases or decreases of activity in other regions. Hence, an inhibitory connection in this case may arise from actual inhibitory projections (e.g. GABAergic), indirect pathways involving intermediary populations, or from withdrawal of excitatory drive. This means ultimately, our interpretations should be understood as functional descriptions of regional connectivity and not claims about specific neurotransmitter systems. Finally, the autonomic testing relied on a brief orthostatic challenge with intermittent blood-pressure readings, which may miss delayed-nOH or other presentations. More detailed clinical testing should be considered in future studies. Notwithstanding, this cohort had relatively mild blood-pressure drops with no significant difference between PD and HC, meaning that the connectivity changes observed are already present in sub-clinically affected individuals, potentially strengthening their relevance as early biomarkers.

## Conclusion

We provide the first *in-vivo* characterization of central autonomic dysfunction in early-PD, which is consistent with known α-synuclein disease pathophysiology. The medullary and hypothalamic connectivity alterations linked to steeper blood-pressure drops, which contrast with the distributed pattern seen in healthy individuals, are consistent with an impaired sympathoexcitatory capacity that predisposes those with PD to falling blood-pressure upon standing (i.e. to nOH). Clinically, the detection of these connectivity changes in a cohort with subclinical autonomic dysfunction suggests that central autonomic affliction precedes overt nOH, and is in-line with longitudinal evidence that sympathoneural dysfunction of the baroreflex identifies those at risk of a central α-synucleinopathy (Goldstein & Sharabi, 2023). Given the strong association between autonomic failure and poor outcomes in PD, including early cognitive dysfunction, these results suggest effect connectivity measures may have the potential to serve as early biomarkers of autonomic vulnerability in PD to help identify and stratify high-risk groups. Longitudinal follow-up of this cohort will now be essential to validate that more pronounced patterns imply worse nOH, and to establish whether these patterns predict the development or worsening of clinically significant nOH.

## Acknowledgements

We would like to thank all the participants who took part in the qMAP-PD study. They donated their time generously and freely to support this work.

